# Domain-specific cognitive impairment 6 months after stroke: the value of early cognitive screening

**DOI:** 10.1101/2023.06.14.23291381

**Authors:** Elise Milosevich, Margaret Moore, Sarah T. Pendlebury, Nele Demeyere

## Abstract

**Background/Objective:** Cognitive screening following stroke is widely recommended, yet few studies have investigated the prognostic value of acute domain-specific function for longer-term cognitive outcome. This study aimed to determine the prevalence of domain-specific impairment acutely and at 6 months, assess the proportion of change in cognitive performance, and examine the predictive value of acute domain-specific cognitive screening.

**Methods:** A prospective cohort of consecutive stroke survivors completed the Oxford Cognitive Screen acutely (≤2 weeks) and 6 months post-stroke. Hierarchical multivariable regression analyses were used to predict general and domain-specific cognitive impairment at 6 months. Demographic/clinical covariates included age, sex, education, atrial fibrillation, hypertension, diabetes, smoking, stroke severity, lesion volume, recurrent stroke, and days to cognitive assessment.

**Results:** A total of 430 stroke survivors (mean age 73.9 years (12.5 *SD*), 46.5% female, median NIHSS 5 [IQR 2-10]) completed 6-month follow-up. Impairments were prevalent within all domains at both timepoints, ranging from 26.7% (n=112) in praxis to 46.8% (n=183) in attention acutely, and 19.6% (n=79) in praxis to 32.6% (n=140) in language at 6 months. Proportion of recovery was highest in praxis (n=73, 71%) and lowest in language (n=89, 46%) and memory (n=82, 48%). Severity of 6-month cognitive impairment was best predicted by the addition of proportion of acute subtests impaired (adjusted *R*^2^=0.298, *p*<0.0001) over demographic/clinical factors alone (adjusted *R*^2^=0.105, *p*<0.0001). Acute cognitive function (β=0.403 SE 0.042, *p*<0.0001) was the strongest predictor of 6-month cognitive performance. Acute domain-specific impairments in memory (β=0.116 SE 0.027, *p*<0.0001), language (β=0.095 SE 0.027, *p*<0.0001) and praxis (β=0.086 SE 0.028, *p*<0.0001) were significant predictors of severity of cognitive impairment at follow-up.

**Conclusion:** Cognitive impairment is highly prevalent initially after stroke across all domains, though impairments in language, memory and attention predominate at 6 months. Early domain-specific screening provides valuable prognostic information with respect to longer-term cognitive functioning.

**Key messages:** *What is already known on this topic:*

- Several demographic, stroke-related, vascular, and brain-related risk factors for post-stroke cognitive impairment have been identified, however, there is a lack of established early domain-specific cognitive markers of long-term cognitive outcome despite an emphasis on routine post-stroke cognitive screening.

*What this study adds:*

- This study showed that severity of acute cognitive impairment identified through early domain-specific screening with the Oxford Cognitive Screen (OCS) was the strongest predictor of cognitive function at follow-up when compared to common post-stroke cognitive risk factors alone. Impairments in memory, language and praxis domains acutely after stroke were particularly important in predicting the severity of cognitive impairment at 6 months.

*How this study might affect research, practice, or policy:*

- This study demonstrated for the first time that early domain-specific screening after stroke with the OCS provides valuable prognostic information with respect to long-term cognitive functioning. Each post-stroke cognitive profile is unique and therefore highlighting different strengths and weaknesses in performance early allows for more accurate information to be communicated to the patient, more tailored discharge care packages and appropriate allocation of rehabilitation resources.

## INTRODUCTION

Cognitive impairment is common following stroke and though prevalence rates vary depending on the nature and timing of assessments, study setting and inclusion criteria, the majority of stroke survivors experience at least one cognitive domain deficit initially after stroke.^1,2^ This is mirrored in subjective reporting, where 90% of 11,000 stroke survivors reported experiencing cognitive problems following stroke.^3^ Identifying which specific cognitive impairments more commonly occur, recover, and persist will aid in intervention planning and in informing stroke survivors and carers when setting realistic expectations and rehabilitation goals.

Early cognitive assessment following stroke is recommended in several clinical stroke guidelines, as well as best practice and expert statements.^4-6^ However, few studies have examined the value of early domain-specific testing. Most research investigating early post-stroke cognitive function to predict longer-term cognitive outcome has used brief global screens,^7^ predominantly the Montreal Cognitive Assessment (MoCA).^8-13^ Early testing with the MoCA was shown to predict long-term cognitive and functional outcome, as well as mortality after stroke.^8^ However, the MoCA is not well suited for many stroke patients as it assumes certain cognitive functions are intact, including speech, reading, writing, spatial attention, motor planning and vision. Consequently, common post-stroke impairments, such as aphasia and hemi-spatial neglect, can contaminate patients’ performance.^7,14^

A detailed neuropsychological battery covering seven domains was used in one previous longitudinal cohort investigating the value of early screening for longer-term cognitive function.^15,16^ Acute domain-specific functioning was found to predict 6-month cognitive and functional outcome better than any other demographic or clinical variable.^16^ Whilst this provides evidence for the utility of acute cognitive testing, a large neuropsychological battery (1.5-2.5 hours) is typically not feasible in an acute setting due to time requirements and increased burden on the patient, as well as staff availability and expertise.

A short domain-specific screen, such as the Oxford Cognitive Screen (OCS),^17,18^ sits between a brief global test and a large detailed neuropsychological battery. The OCS does not rely on intact language function, is feasible for stroke patients with aphasia, apraxia, and hemi-spatial neglect, and has been shown to be more sensitive in detecting impairments than the Mini-Mental State Examination (MMSE) and the MoCA.^1,14^ It has also been demonstrated to reliably tap established anatomical patterns of functional specialization.^19^ The OCS is currently used routinely within acute stroke clinical settings for post-stroke cognitive screening, however its prognostic value in determining longer-term cognitive outcome has yet to be investigated. Therefore, this study aimed to (i) determine the prevalence of domain-specific cognitive impairments acutely and at 6 months post-stroke, (ii) assess the proportion of change in domain-specific function from acute to follow-up, as well as (iii) examine the predictive value of early domain-specific screening using the OCS with respect to cognitive function at 6-months.

## METHODS

### Standard protocol approvals, registrations, and patient consents

The present study considered existing data from a cohort of acute stroke survivors recruited through the OCS-Tablet and OCS-Recovery studies (ethical approval by the National Research Ethics Committee (UK) references 14/LO/0648 and 18/SC/0550, respectively). Acute stroke patients were included if they were aged ≥18 years, able to give written or witnessed informed consent, concentrate for 20 minutes, and had sufficient English language comprehension. All procedures were in accordance with the Declaration of Helsinki.

### Participants

A consecutive sample of acute stroke patients was recruited through the Oxford Cognitive Screening programme based within the John Radcliffe Hospital, UK, acute stroke unit, from 2012-2019. A total of 866 stroke patients were recruited and assessed acutely (≤2 weeks), 430 (49.7%) of whom completed 6-month follow-up (Figure 1).

**Figure 1.**
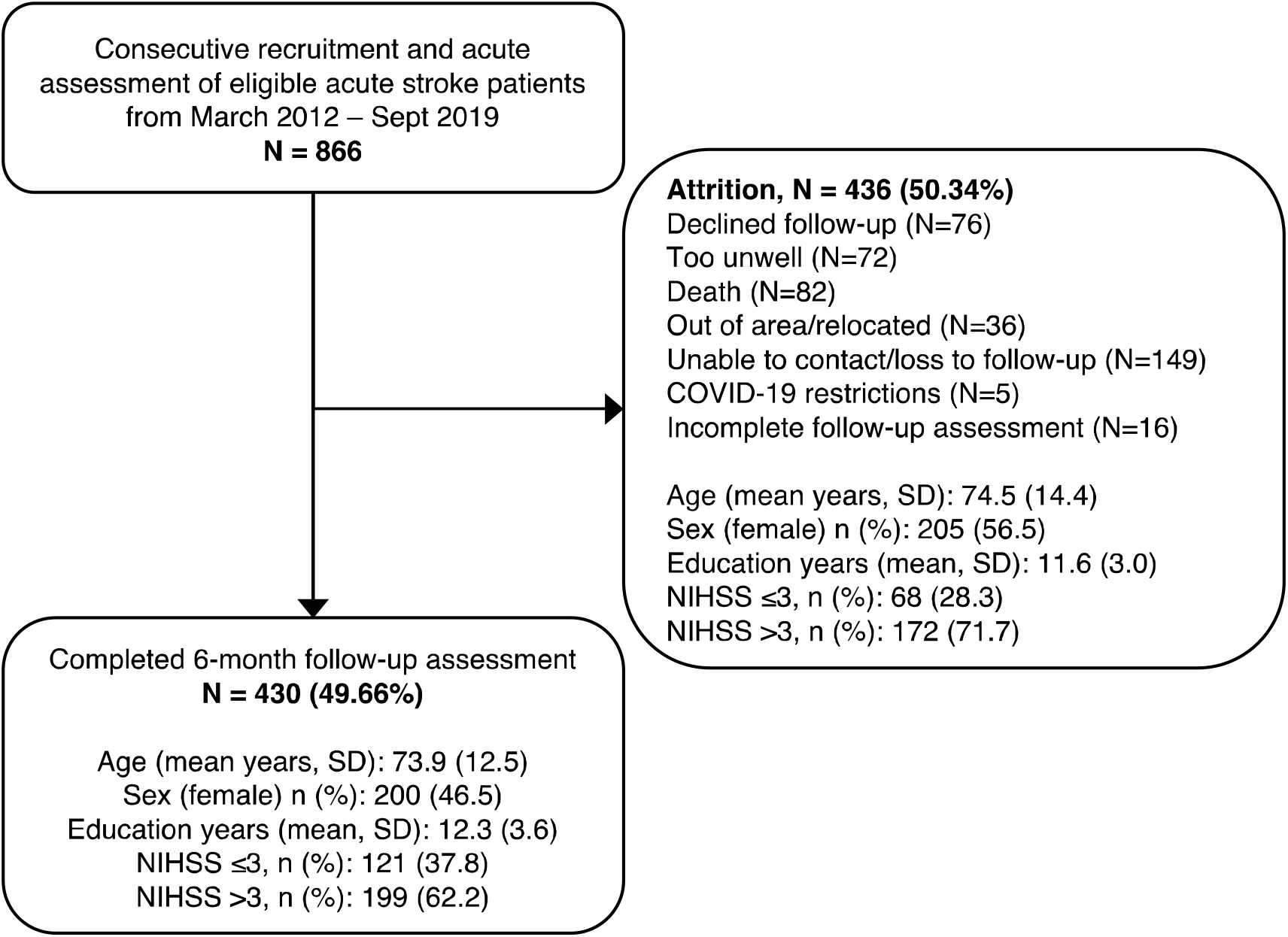
Flow chart of patient cohort from baseline to 6-month follow-up.

### Cognitive assessment

Domain-specific assessments acutely and at follow-up were carried using the Oxford Cognitive Screen (OCS).^17^ Twelve subtest scores were categorized into 6 cognitive domains: language (picture naming, semantic understanding, sentence reading), attention (egocentric and sustained attention, allocentric attention), executive function (trail-making), memory (orientation, verbal memory, episodic memory), praxis (gesture imitation), and number processing (calculation and number writing). Subtests were binarized into impaired or unimpaired based on normative scores for each subtest.^17,18^ A domain impairment was characterized as at least one impaired subtest in that domain as number of subtests range from 1-3 across domains. The OCS was administered by trained neuropsychologists and occupational therapists at both bedside acutely and at in-person follow-up assessments (supplementary Methods).

### Statistical analysis

Preliminary analyses examined whether any systematic differences in clinical characteristics or cognitive function were present between patients who did and did not complete follow-up (*t*-tests for continuous data and χ^2^ test for categorical data). Descriptive statistics were used to determine the prevalence and nature of cognitive impairment and recovery. The degree of association between paired domain and subtest impairments acutely, at 6 months, and from acute to follow-up, was determined using tetrachoric correlation analysis.^20^ This was repeated using receiver operating curve (ROC) analysis to clarify the sensitivity/specificity of acute impairments for prediction of 6-month domain-specific outcome.

Hierarchical multivariable regression analyses were used to determine the value of early cognitive screening for predicting longer-term cognitive outcome. Three models were produced progressing from least to most in cognitive detail. The first model examined the relationship between proportion of cognitive subtests impaired acutely and at follow-up. The second examined acute domain-specific function (impaired/unimpaired) and proportion of subtests impaired at follow-up. The third model examined each of the 6 domains individually (impaired/unimpaired) at acute and follow-up. Demographic/clinical factors were entered in the first block, followed by the addition of acute cognition in the second block, with cognitive performance at follow-up considered as the dependent variable. The general rule of at least 10 events required per candidate predictor parameter (EPP) was applied.^21^ Demographic covariates included age, sex, and years of education. Clinical variables included stroke severity (NIHSS), lesion volume, recurrent stroke, atrial fibrillation, hypertension, diabetes, smoking, and days from stroke onset to cognitive assessment. Scans and lesion masks were pre-processed in line with the standard processing protocol reported by Moore and Demeyere.^19^ Lesion volume was calculated by summing the total of number of lesioned voxels and converting this value into square centimetres. Missing NIHSS data was amended through multiple imputation, though initial sensitivity analyses were completed using complete NIHSS data and no significant difference was found. Core analyses were repeated within the subset of patients with first-ever stroke. For all analyses, *p* values <0.05 were considered statistically significant. Bonferroni correction was applied due to the increased risk of type I error through multiple comparisons. All analyses were performed in R version 4.0.5.

### RESULTS

Of the 866 patients assessed acutely, 430 (49.7%) completed follow-up assessment at 6 months (Figure 1). Patient demographics did not statistically differ between those who were and were not re-assessed (supplemental Table 1). Stroke severity did differ between groups (χ^2^=5.096, p=0.002), with a greater proportion more severe stroke (NIHSS >3) (71.7%) occurring in the attrition group compared to those followed-up (62.2%). Additionally, the attrition group demonstrated a significantly (p<0.001) greater proportion of impairment in language (58.3% vs 45.2%), executive function (36.4% vs 29.3%), memory (56.0% vs 39.9%), number (49.0% vs 41.7%) and praxis (39.1% vs 26.7%) (see supplemental Table 2). Notably however, a greater proportion of patients with no cognitive impairment were also not re-examined (14.5% vs 1.6% in those with follow-up data), suggesting attrition occurred both in cases of more severe stroke and in those who had no acute cognitive impairments.

The mean age of those who completed follow-up was 73.86 years (12.51 SD), 46.5% were female, and the mean years of formal education received was 12.25 (3.55 SD). Demographics and acute clinical characteristics of the cohort is outlined in Table 1. Cognition was assessed with the OCS acutely at a mean of 4.39 (4.46 SD) days after stroke and again at a mean 6.65 (1.06 SD) months.

**Table 1.** Cohort demographics and acute clinical characteristics.

|  | **N = 430** |
| --- | --- |
| Age at stroke, mean (SD) | 73.86 (12.51) |
| Sex (female), n (%) | 200 (46.51) |
| Education years, mean (SD) | 12.25 (3.55) |
| Handedness, right n (%) | 374 (86.98) |
| *Stroke subtype, n (%)* |  |
| Ischemic | 362 (84.19) |
| Haemorrhagic | 65 (15.12) |
| Mixed | 3 (1.00) |
| *Lesion side, n (%)* |  |
| Left | 153 (35.58) |
| Right | 168 (39.07) |
| Bilateral | 34 (7.91) |
| Undetermined | 75 (17.44) |
| *Major vascular territory, n (%)*<sup>A</sup> |  |
| Anterior | 39 (9.07) |
| Middle | 177 (41.16) |
| Posterior | 59 (13.72) |
| Vertebrobasilar | 55 (12.79) |
| Multifocal | 8 (1.86) |
| Lacunar | 18 (4.19) |
| Undetermined | 74 (17.21) |
| First-ever stroke, n (%) | 292 (67.91) |
| NIHSS, median [IQR]<sup>B</sup> | 5 [2–10] |
| NIHSS ≤3, n (%) | 121 (37.81) |
| NIHSS >3, n (%) | 199 (62.19) |
| Modified Rankin Scale, median [IQR]<sup>C</sup> | 1 [0–2] |
| Barthel Index, median [IQR]<sup>C</sup> | 15 [9–19] |
| Independent at admission, n (%) | 376 (87.44) |
| Dependent (care required), n (%) | 54 (12.56) |
| *Comorbidities, n (%)* |  |
| CCI low (0 – 1) | 268 (62.33) |
| CCI high (≥2) | 162 (37.67) |
| Atrial fibrillation | 109 (25.35) |
| Hypertension | 261 (60.70) |
| Diabetes mellitus | 83 (19.30) |
| *Smoking, n (%)* |  |
| Current | 44 (10.23) |
| Past | 34 (7.91) |
| Never | 352 (81.86) |
| Days from stroke to T1 assessment, mean (SD) | 4.38 (4.46) |
| Length of stay in hospital, mean days (SD) | 11.64 (11.44) |

CCI: Charlson Comorbidity Index; NIHSS: National Institute of Health Stroke Scale (≤3 minor, >3 major) Demographics/clinical characteristics were determined on admission; CCI was scored based on the ICD-10 scoring scheme through medical records at hospital discharge. Ethnicity was not formally documented, however based on population-based data inclusive of these patients (Oxford Vascular Study<sup>43</sup>) the presumed predominant ethnicity (>90%) was White-British.
<sup>A</sup> Note that vascular supply is a crude characterization of primary territory affected
<sup>B</sup> NIHSS recorded for n=320 (74%) due to not being routinely recorded in this clinical setting prior to 2014
<sup>C</sup> Pre-morbid modified Rankin Scale (mRS) reported for n=248 (58%)
<sup>D</sup> Barthel Index was reported for n=141 (33%)
<sup>E</sup> Dependence was categorized as requiring formal support (by family n=18, by carer n=36)

### Prevalence of and associations between domain-specific impairments acutely and at 6 months

Prevalence of post-stroke cognitive impairments acutely and at 6 months is outlined in Table 2. Of the 430 participants, 423 (98.4%) experienced at least one subtest impairment within a cognitive domain acutely (n=316; 73.7% exhibited multi-domain deficits), and 293 (68.1%) were impaired at 6 months (n=197; 45.8% multi-domain). Impairments on at least one subtest were prevalent across all domains both acutely and at 6-months, ranging from 112 (26.7%) impaired in praxis to 183 (46.8%) in attention acutely, and 79 (19.6%) in praxis to 140 (32.6%) in language at 6 months (Figure 2). Associations between paired domain impairments at acute and 6 months is shown in Figure 3. The strongest associations were found between impairments in memory and number processing (acute *r*_tet_=0.69, p<0.001; 6 months *r*_tet_=0.58, p<0.001), as well as language and number processing (acute *r*_tet_=0.62, p<0.001; 6 months *r*_tet_=0.56, p<0.01) (Figure 3).

**Table 2.** Prevalence of post-stroke cognitive impairment acutely and at 6-month follow-up.

| Domain | Assessed acute (N) | Acute impaired N (%) | Assessed follow-up N | Follow-up impaired N (%) |
| --- | --- | --- | --- | --- |
| **Language** | **429** | **194 (45.2)** | **430** | **140 (32.6)** |
| Picture naming | 429 | 138 (32.2) | 428 | 78 (18.2) |
| Semantic understanding | 427 | 43 (10.1) | 412 | 16 (3.9) |
| Sentence reading | 419 | 136 (32.5) | 422 | 88 (20.9) |
| **Spatial attention** | **391** | **183 (46.8)** | **415** | **129 (31.1)** |
| Egocentric attention | 391 | 124 (31.7) | 415 | 57 (13.7) |
| Allocentric attention | 391 | 108 (27.6) | 414 | 101 (24.4) |
| **Executive Function** | **379** | **111 (29.3)** | **405** | **98 (24.2)** |
| **Memory** | **429** | **171 (39.9)** | **430** | **137 (31.9)** |
| Orientation | 427 | 90 (21.1) | 426 | 72 (16.9) |
| Verbal memory | 427 | 107 (25.1) | 417 | 78 (18.7) |
| Episodic memory | 391 | 79 (20.2) | 416 | 53 (12.7) |
| **Number processing** | **427** | **178 (41.7)** | **418** | **85 (20.3)** |
| Calculations | 427 | 73 (17.1) | 417 | 39 (9.4) |
| Writing | 411 | 160 (38.9) | 407 | 64 (15.7) |
| **Praxis** | **419** | **112 (26.7)** | **403** | **79 (19.6)** |
| **Any domain** | 430 | 423 (98.4) | 430 | 293 (68.1) |
| **Multi-domain** | 429 | 316 (73.7) | 430 | 197 (45.8) |
| **Single domain** | 430 | 107 (24.9) | 430 | 96 (22.3) |

Prevalence of impairment within each domain at both acute assessment and at 6-months follow-up. See eTable 2 for a summary of proportions of patients who were not assessed at each time point. Any domain impairment was defined by an impairment on any subtest in any domain; multi-domain refers to impairment in more than one domain.

**Figure 2.**
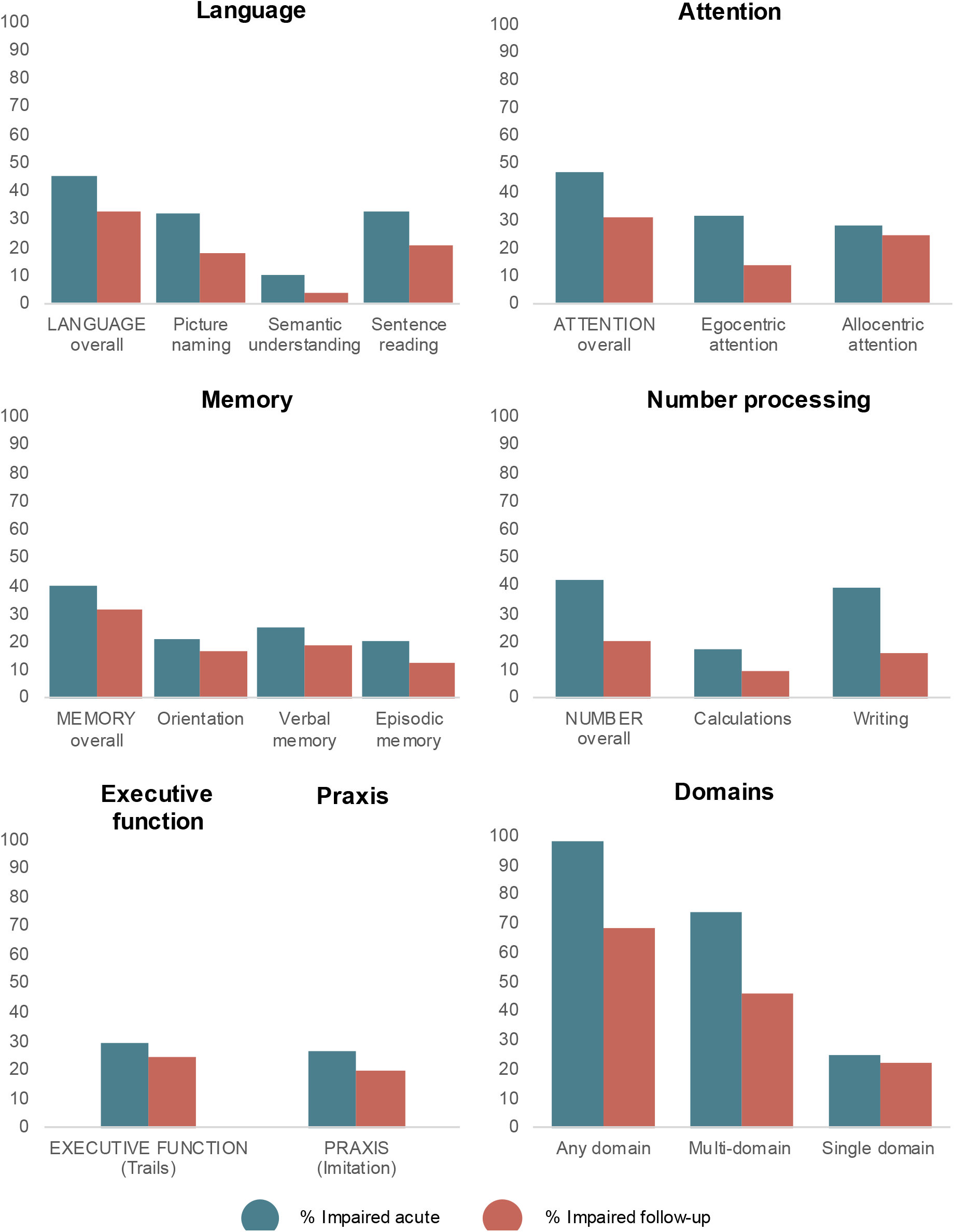
Prevalence of domain-specific impairment acutely and at 6-month follow-up. Each box contains prevalence of impairment in each domain overall, as well as by each subtask pertaining to each domain. Executive function and praxis are in the same box for visual purposes only. ‘Any domain’ impairment refers to impairment in any domain, ‘multi-domain’ is impairment in more than one domain, and ‘single domain’ is impaired in only one domain.

**Figure 3.**
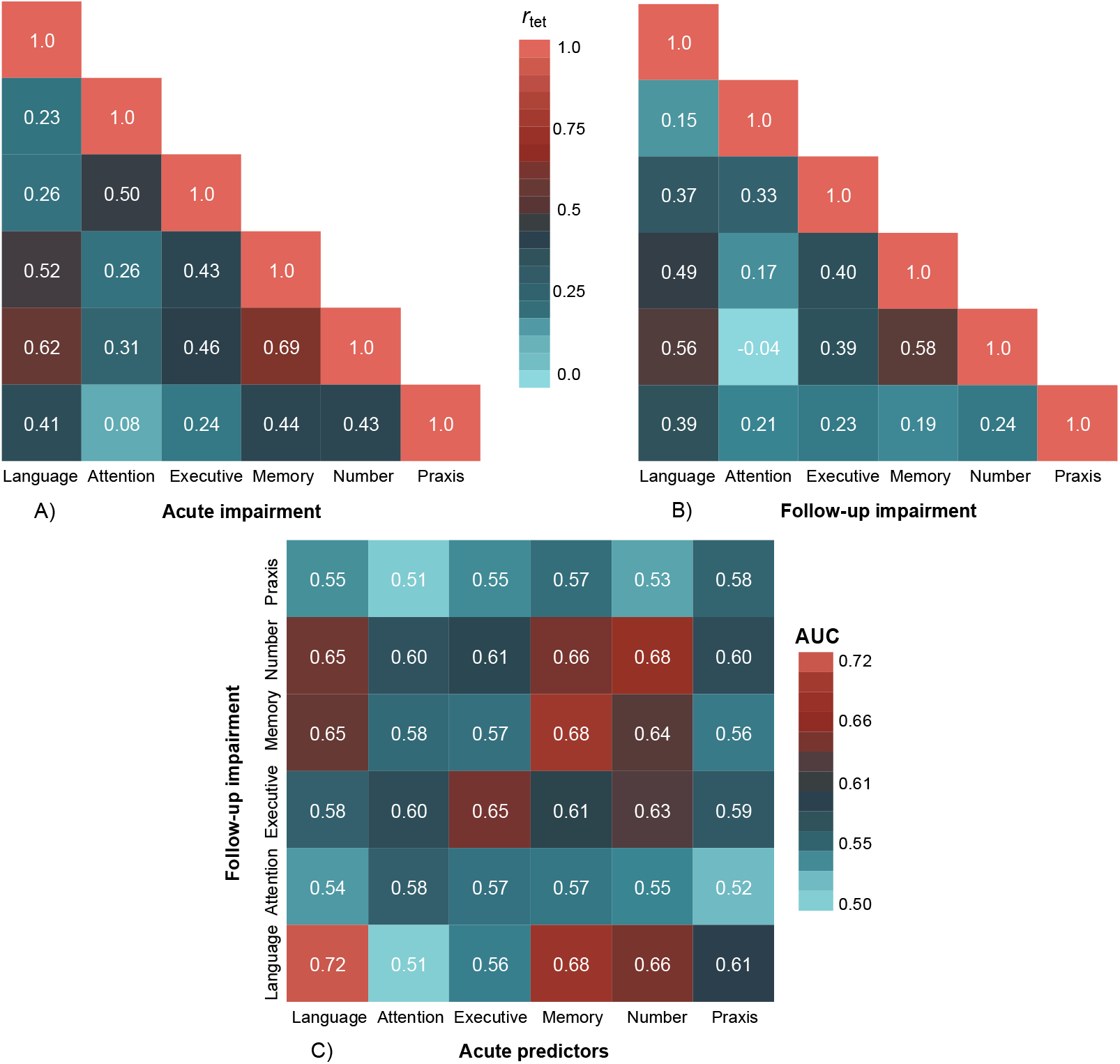
Association between domain impairments at acute assessment and at follow-up, as well as acute impairments predictive of follow-up impairments. **A)** Associations between acute domain impairments. **B)** Associations between 6-month follow-up impairments. **C)** Acute impairments predictive of 6-month follow-up impairments. Both ***A*** and ***B***: Tetrachoric correlation coefficient is shown in each box with a gradient extending from 0 (light blue, no association) to 1 (light red, complete association) for domain-specific impairments. ***C***: Area under the Receiver Operating Characteristic curve (AUC) values are presented in each box with a gradient extending from 0 (light blue, completely inaccurate prediction) to 1 (light red, complete prediction), acute: x-axis, follow-up: y-axis.

### Proportion of change in cognitive function from acute to 6 months

Prevalence of impairment on at least one subtest decreased across all domains from acute to 6 months (Figure 2; Figure 4). Proportion of recovered impairments was highest in praxis (70.9%), followed by number processing (65.3%), executive function (57.5%), attention (56.2%), memory (48.0%) and language (45.9%) (Figure 5; supplementary Table 3). Most participants who were unimpaired acutely remained unimpaired at follow-up (Figure 4; Figure 5). However, newly acquired impairments were found on subtests in each domain, predominantly in attention (19.6%) and memory (18.6%) (Figure 5). The strongest associations between impairments from acute to 6 months was between language (both timepoints) (*r*_tet_=0.56, p<0.01), memory (acute) and language (follow-up) (*r*_tet_=0.55, p<0.001), and memory (both timepoints) (*r*_tet_=0.54, p<0.001. Similar effects were present in ROC analysis. Specifically, highest AUC values were present for acute and 6-month language impairments (AUC=0.72), acute and 6-month memory impairments (AUC=0.68), acute memory predicting 6-month language (AUC=0.68), and for acute and 6-month number impairments (AUC=0.68) (Figure 3c). The degree of relation between domain subtests acutely and at 6 months, and from acute to 6 months is shown in supplementary Figure 1.

**Figure 4.**
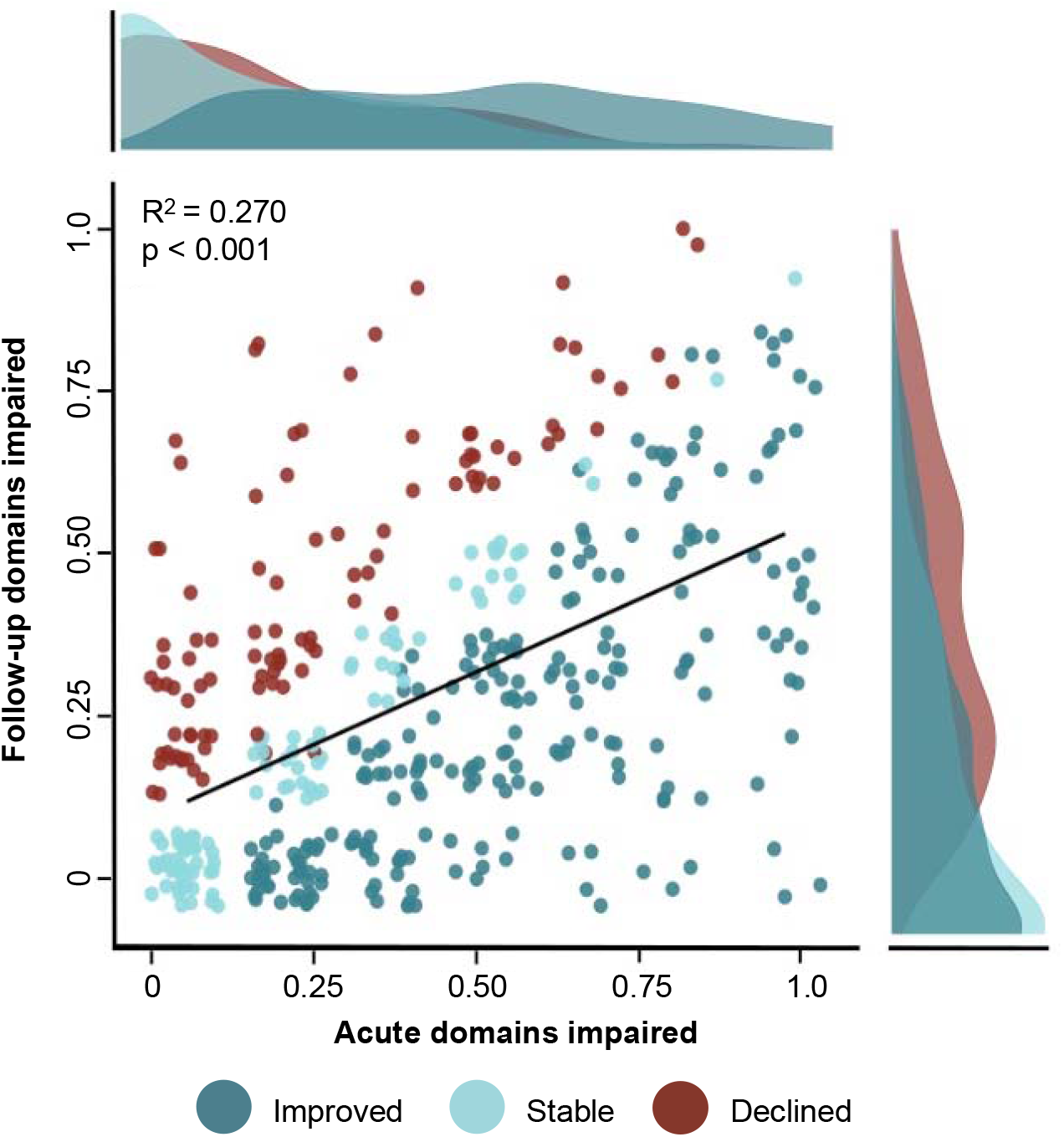
Proportion of change across cognitive domains from acute to 6 months. Improved (dark blue) is characterized by a reduction in proportion of impaired domains, stable (light blue) shows those who remained impaired in the same proportion of domains and declined (red) signifies those who increased in proportion of impaired domains. Solid trend line represents the reported regression analysis with the considered clinical/demographic covariates accounting for 27% of variance.

**Figure 5.**
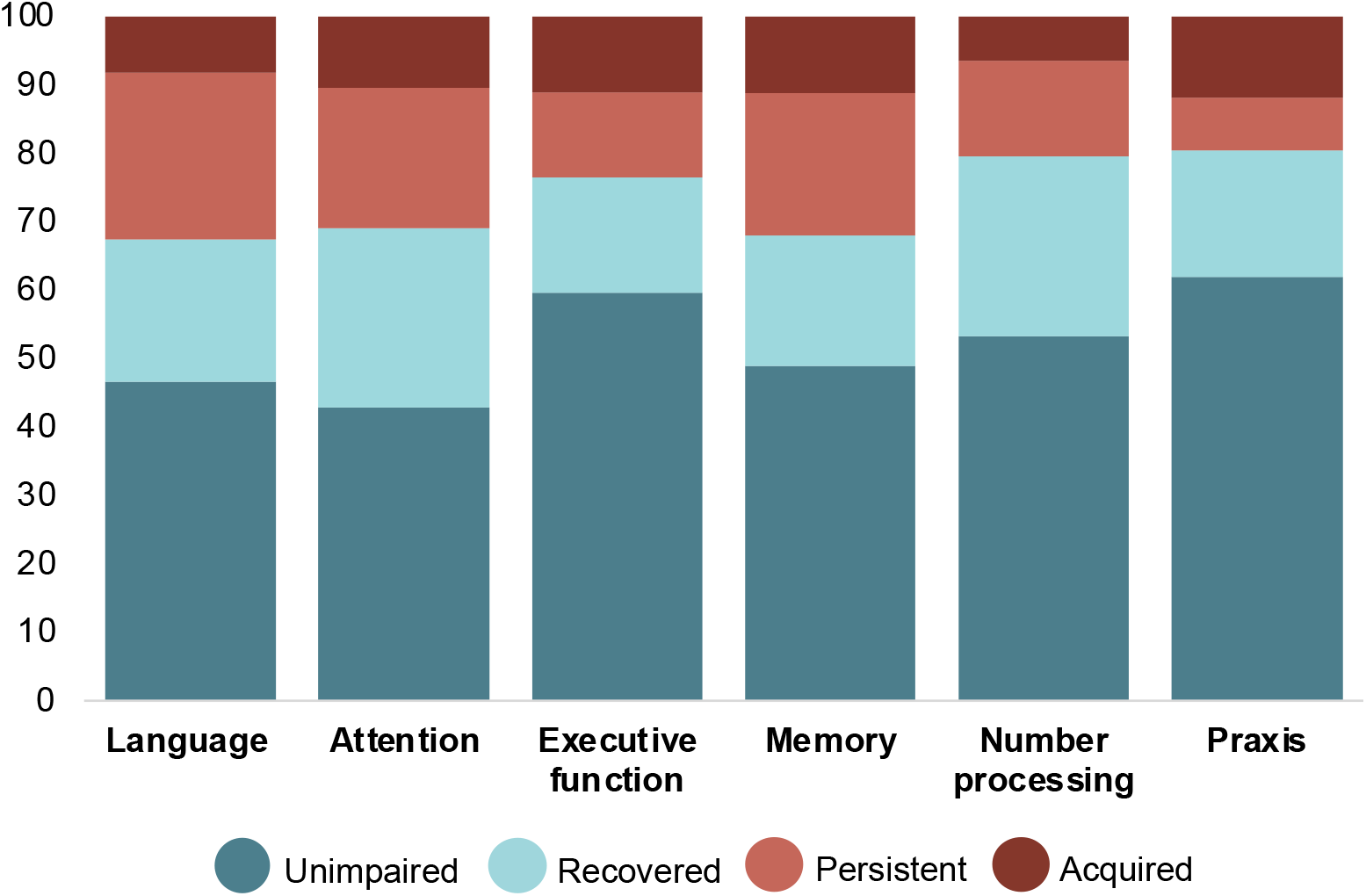
Proportion of change within each cognitive domain from acute to 6 months. Unimpaired (dark blue) refers to proportion of individuals who remained unimpaired, recovered (light blue) refers to those who were impaired, but were no longer impaired at follow-up, persistent (light red) refers to individuals who remained impaired and acquired (red) refers to those who were unimpaired, but gained a new impairment at follow-up assessment. On average across all domains, 52.2% remained unimpaired, 21.3% recovered, 16.6% had persistent impairment, and 9.9% acquired a new impairment.

### Predictors of proportion of impaired cognitive domain subtests at 6 months

Hierarchical regression showed the base model (Block 1) of common risk factors to significantly predict proportion of subtests impaired at follow up (*F*(11,333)=4.69, *p*<0.0001, adjusted *R*^2^=0.105) (Table 3). In this model, age (β=0.005, *p*<0.0001), education (β=-0.009, *p*=0.012), smoking (β=0.072, *p*=0.026) and lesion volume (β=0.000, *p*=0.004) were significantly associated, though only age and lesion volume remained significant after correction (Bonferroni corrected alpha level=0.005). Proportion of acute subtests impaired was then added to this base model of clinical factors (Model 1, Block 2), which improved the model, adjusted *R*^2^=0.298 (*p*<0.0001). After correction, only age (β=0.004, *p*<0.0001) and acute cognition (proportion of impaired subtests acutely) (β=0.403, *p*<0.0001) remained significant. In Model 2 (Block 2) acute domain-specific cognitive impairments were added to the base model, which explained slightly more variance, adjusted *R*^2^=0.309 (*p*<0.0001) (Table 3). In this expanded model, acute language (β=0.095, *p*=0.0002), memory (β=0.116, *p*<0.0001) and praxis impairments (β=0.084, *p*=0.003), as well as age (β=0.004, *p*<0.0001) remained significant after correction. Notably, all hierarchical regression analyses yielded similar results within the subset of patients with first-ever stroke (see supplementary Analysis).

**Table 3.** Results of regression analyses for clinical/demographic factors and acute cognitive impairment predictive of cognitive impairment at 6 months (proportion of subtests impaired).

|  | Proportion of OCS subtests impaired at 6 months |  |  |
| --- | --- | --- | --- |
| **Block 1: Demographic/Clinical factors** | $\beta$ (SE) | *t* value | *p* value |
| Age | 0.094 (0.001) | 4.817 | 0.000*<sup>a</sup> |
| Sex (male), | -0.006 (0.026) | -0.222 | 0.825 |
| Education years | -0.009 (0.004) | -2.527 | 0.012* |
| Atrial fibrillation | -0.025 (0.030) | -0.847 | 0.398 |
| Hypertension | -0.031 (0.026) | -1.182 | 0.238 |
| Diabetes | 0.041 (0.031) | 1.325 | 0.186 |
| Smoking | 0.073 (0.032) | 2.241 | 0.026* |
| NIHSS | 0.004 (0.003) | 1.372 | 0.171 |
| Recurrent stroke | 0.045(0.032) | 1.646 | 0.107 |
| Days to assessment | 0.001 (0.003) | 0.476 | 0.635 |
| Lesion Volume | <0.001 (<0.001) | 2.923 | 0.004* |
| **R<sup>2</sup> = 0.134, Adjusted R<sup>2</sup> = 0.105, F = 4.693, 11 and 333 df, *p* <0.0001** |  |  |  |
| **Block 2 (Model 1):**<br>**Proportion of acute subtests impaired** | $\beta$ (SE) | *t* value | *p* value |
| Age | 0.005 (0.001) | 4.674 | 0.000*<sup>a</sup> |
| Sex (male) | -0.004 (0.023) | -0.197 | 0.844 |
| Education years | -0.005 (0.003) | -1.527 | 0.128 |
| Atrial fibrillation | -0.032 (0.026) | -1.223 | 0.222 |
| Hypertension | -0.033 (0.023) | -1.452 | 0.147 |
| Diabetes | -0.006 (0.028) | -0.213 | 0.831 |
| Smoking | 0.006 (0.029) | 2.066 | 0.040* |
| NIHSS | -0.000 (0.002) | -0.134 | 0.893 |
| Recurrent stroke | 0.045 (0.024) | 1.862 | 0.064 |
| Days to assessment | 0.002 (0.003) | 0.786 | 0.433 |
| Lesion Volume | <0.001 (<0.001) | 1.132 | 0.2585 |
| **Severity of acute cognitive impairment** | 0.403 (0.042) | 9.616 | 0.000*<sup>a</sup> |
| **R<sup>2</sup> = 0.323, Adjusted R<sup>2</sup> = 0.298, F = 13.190, 12 and 332 df, *p* <0.0001** |  |  |  |
| **Block 2 (Model 2):**<br>**Acute domain-specific impairment** | $\beta$ (SE) | *t* value | *p* value |
| Age | 0.004 (0.001) | 4.266 | 0.000*<sup>a</sup> |
| Sex (male) | 0.009 (0.023) | 0.399 | 0.690 |
| Education years | -0.005 (0.003) | -1.475 | 0.141 |
| Atrial fibrillation | -0.027 (0.026) | -1.040 | 0.299 |
| Hypertension | -0.027 (0.023) | -1.182 | 0.238 |
| Diabetes | -0.003 (0.029) | -0.094 | 0.925 |
| Smoking | 0.076 (0.029) | 2.618 | 0.009* |
| NIHSS | 0.001 (0.002) | 0.529 | 0.597 |
| Recurrent stroke | 0.039 (0.024) | 1.619 | 0.107 |
| Days to assessment | 0.000 (0.003) | 0.155 | 0.877 |
| Lesion Volume | 0.000 (0.000) | 0.715 | 0.475 |
| **Acute Language** | 0.095 (0.027) | 4.303 | 0.000*<sup>a</sup> |
| **Acute Attention** | 0.011 (0.024) | 0.446 | 0.656 |
| **Acute Executive** | 0.065 (0.027) | 2.376 | 0.018* |
| **Acute Memory** | 0.116 (0.027) | 4.303 | 0.000*<sup>a</sup> |
| **Acute Number** | 0.041 (0.028) | 1.477 | 0.141 |
| **Acute Praxis** | 0.084 (0.028) | 3.042 | 0.002*<sup>a</sup> |
| **R<sup>2</sup> = 0.345, Adjusted R<sup>2</sup> = 0.309, F = 9.612, 167 and 310 df, *p* <0.0001** |  |  |  |
| * Significance *p* <0.05 |  |  |  |
| <sup>a</sup> Remained significant after Bonferroni correction for multiple comparisons |  |  |  |

### Predictors of domain-specific cognition at 6 months

A series of 6 hierarchical regression analyses (supplemental Tables 4-10) were conducted to identify acute factors predictive of individual domain impairments at follow-up. For each of these models, a base model reported how well conventional predictors explained the occurrence of domain-specific impairment at 6 months, while an expanded model investigated how the addition of acute domain-specific function improved this fit. With the addition of all acute domain impairments, the language model improved from AIC 222.9 (p=0.3792) to AIC 196.02 (*p*=0.0004); strongest predictor of a language impairment was acute language (OR=2.827, *p*=0.0149) and memory (OR=2.6071, *p=*0.0388) (supplemental Table 5). Attention model improved slightly from AIC 236.3 (p=0.0377) to AIC 235.3 (*p*=0.0737); strongest predictor was age (OR=1.0673, *p*=0.0006) (supplemental Table 6). Executive function model improved from AIC 193.44 (p=0.0231) to AIC 170.1 (*p*<0.0001); strongest predictors were acute executive dysfunction (OR=6.266, *p*=0.0005), age (OR=1.078, *p*=0.003) and sex (OR=3.982, *p*=0.006) (supplemental Table 7). The model predicting memory improved from AIC 221.4 (p=0.379) to AIC 196.0 (*p*=0.0004); strongest predictors were acute memory (OR=2.607, *p*=0.0388) and language (OR=2.826, *p*=0.0149) (supplemental Table 8). Number processing model improved from AIC 166.2 (p=0.154) to AIC 157.3 (*p*=0.0075); strongest predictors were acute praxis (OR=2.7268, *p*=0.0809), smoking (OR=2.6746, p=0.0674) and age (OR=0.9427, *p*=0.0209), however none remained significant after correction (supplemental Table 9). Lastly, the praxis model improved from AIC 209.3 (p=0.2818) to AIC 202.9 (*p*=0.0746); strongest predictor was acute praxis impairment (OR=3.2656, *p*=0.0079) (supplemental Table 10).

## DISCUSSION

In this cohort of 430 stroke survivors, nearly every patient was impaired on at least one domain subtest initially following stroke, while just over two-thirds were impaired at 6 months, many of which exhibited multi-domain deficits. Subtest impairments within attention and language domains were most prevalent acutely with the addition of impairments in memory at 6 months. The prevalence of impairment decreased within all domains from acute to 6 months, and most participants who were unimpaired acutely remained unimpaired at follow-up. However, newly acquired impairments were found across all domains at 6 months. The severity of acute domain-specific cognitive impairment identified through early cognitive screening with the OCS was shown to be the strongest predictor of cognitive function at follow-up when compared to common post-stroke cognitive risk factors alone. Acute impairments in memory, language and praxis domains were particularly important in predicting the severity of cognitive impairment at follow-up. Each domain-specific impairment at follow-up was best predicted by the same domain impairment acutely except attention (better predicted by age) and number impairments, where no predictor remained significant after correction.

The high prevalence of impairment is in line with previous studies reporting overall occurrence of cognitive impairment initially after stroke (ranging from 49%–92%)^1,9,22-25^ and at 6 months (41%–57%).^25-27^ Our findings of slightly higher prevalence at both timepoints are likely due to the methodology concerning the way in which cognition was defined and the recruitment of an inclusive sample of patients with potential pre-stroke cognitive decline, recurrent strokes, and individuals with severe aphasia who are often excluded from research.^28^ Additionally, the OCS has been shown to have higher sensitivity in detecting stroke-specific deficits compared to frequently used dementia-based global cognitive screening tools.^1,14^ The increased prevalence of multi-domain impairments found in this cohort supports previous findings by Nys et al.,^15^ who reported that impaired patients experienced an average of 3 domain deficits. Similarly, Jokinen et al.,^29^ found 83% were impaired in at least one domain and 50% were impaired in 3 or more 3 months post-stroke. The frequency of multi-domain impairments is reflected in clinical guidance for stroke that states each cognitive domain should not be considered in isolation,^4^ emphasizing the need for multi-domain screening. Furthermore, brief global screens that are primarily focused on verbal memory and fail to assess stroke-specific deficits, such as spatial neglect and core language abilities (reading or writing), may omit key information that can be invaluable to rehabilitation and discharge planning.^1,14^

The frequency of domain-specific deficits reported by previous studies also varies. For example, both Hurford et al.,^24^ and Leśniak et al.,^23^ found attention to be the most commonly affected domain, though their attention measures included different executive control demands. Different cognitive measures and how domains are characterized may partly explain the variability across studies. In our cohort, attention and language were the most frequently impaired domains, which are hallmark deficits of lateralized stroke.^30,31^ A high prevalence of spatial attention impairments was also found at 3 months in previous studies.^29^ Both egocentric and allocentric neglect were prevalent in both left and right-sided stroke, highlighting that allocentric neglect and right-sided neglect are not as uncommon as previously thought.^32^ High rates of language impairment in this sample may be explained by aphasia not being a study exclusion criterion in effort to capture the true spectrum of post-stroke deficits. A meta-analysis concluded aphasia is present in approximately 30% of acute stroke patients,^33^ therefore excluding aphasic patients markedly biases post-stroke cognitive profiles. Additionally, though number processing, memory, executive function, and praxis deficits were less frequent in this study, impaired subtests still occurred in 27-42% acutely and 20-32% at 6 months. This is consistent with previous studies highlighting executive function, processing speed and episodic memory as more commonly reported deficits months after stroke.^22,24,29^ Overall, it is clear cognitive impairments occur in multiple domains, frequently affecting complex cognitive abilities in which attention and language have a major role.

The prevalence of impairment decreased across all domains from acute to follow-up, which aligns with previous studies.^15,16,24^ The highest proportion of recovery was observed within praxis and number processing domains, while language and memory impairments were most persistent. This was similar to the findings of Hurford et al.,^24^ where memory deficits did not significantly change between assessments but contradicted the results of Turunen et al.,^25^ which found the greatest rates of recovery from baseline to 6 months in executive functions and visual memory. Persistent and acquired memory deficits could be due to memory typically being associated with pre-stroke neurodegeneration, which would not be expected to change over a relatively short period. Similarly, persistent language deficits were consistent with reports of long-term language impairments at one-year post-stroke,^34^ and a meta-analysis which found cases of language recovery to decrease beyond 6 months post-stroke.^35^ Another important finding is the proportion of acquired impairments observed within all domains, ranging from

11%–20, and consistent with Nys et al.,^16^ who reported new impairments at 6-month follow-up in all five domains assessed. The presence of varying domain outcomes in this sample mirrors previous varying overall trends towards recovery, stability and decline.^36,37^ In general, the risk of post-stroke cognitive decline is dependent on the combination of pre-existing cerebral vulnerability/reserve and the impact of the stroke lesion.^38^

The key finding of this study is that routine early domain-specific screening with the OCS can be used to predict 6-month cognitive function and provide domain-specific profiling, which can inform tailored neurorehabilitation strategies. The overall aim of this study was not to identify an optimal predictive model, but to determine if acute cognition could provide useful prognostic information. Acute cognitive functioning was strongly associated with both severity of cognitive impairment at follow-up and domain-specific impairment, explaining approximately 30% more variance in the outcomes than conventional demographic and clinical factors alone. More specifically, memory, language and praxis deficits acutely were highly significant predictors of the severity of cognitive impairment at follow-up. Age emerged as the most consistently significant demographic risk factor for severity of cognitive impairment at 6 months and was the strongest predictor of a 6-month attention impairment. Collectively, these findings indicate that both age and acute domain-specific cognitive function as measured by the OCS are important when considering neurorehabilitation guidance and possible cognitive trajectories 6-months post-stroke. These findings support the current recommendation for routine early domain-specific cognitive screening post-stroke and demonstrates the prognostic value of early cognitive markers, both of which have been highlighted as priorities for post-stroke cognitive impairment research by the European Stroke Organisation and European Academy of Neurology, American Heart Association, and Canadian Best Practice.^5,39,40^

This study has strengths, including the consecutive recruitment of an inclusive sample with moderate and severe aphasia, a balance of both minor and major stroke, as well as longitudinal domain-specific cognitive screening. However, there are also limitations. Though efforts were made to be as inclusive as possible, the nature of cognitive studies requiring a sufficient level of comprehension in order to consent may preclude those with most severe stroke. All domains were assessed for each individual, though a minority of patients were not able to complete all domain subtests, which may cause some bias and under representation of cases with the poorest cognitive outcome. Also, patients who were not re-examined at 6 months demonstrated a significantly greater proportion of impairment in almost all domains acutely, which could indicate a level of attrition bias. However, there was also a greater proportion of unimpaired patients in the cohort that was not re-examined. A substantial proportion of the attrition was due to death (19%), which represents another potential source of bias towards underrepresentation of more severe stroke and severe cognitive impairment at follow-up, as well as multi-domain impairments. Although, attrition in a representative stroke sample is generally unavoidable and a relatively large sample size was retained with moderate stroke on average (median NIHSS 5, range 0-30). Lastly, though the OCS has been shown to be highly sensitive, it may lack specificity similar to the MoCA, which could result in overestimation of impairment.

In summary, early domain-specific cognitive profiling with the OCS provides valuable prognostic information with respect to longer-term cognitive functioning. The OCS is currently used routinely in clinical settings and these findings suggest it could be employed for prognostic purposes within a stroke care pathway, prompting further follow-up with a more detailed neuropsychological battery. Cognitive impairment was shown to be highly prevalent initially after stroke, though after 6 months, prevalence of impairment had decreased across all domains demonstrating a general trend toward recovery. However, persistent impairments were also found across all domains, particularly memory and language deficits. Each post-stroke cognitive profile is unique and therefore highlighting different strengths and weaknesses in performance early allows for more accurate information to be communicated to the patient, more tailored discharge care packages and appropriate allocation of rehabilitation resources.

## Supporting information

Strobe Checklist Cohort Studies

## Data Availability

All data produced in the present study are available upon reasonable request to the authors.

## ACKNOWLEDGMENTS

We would like to thank Ms Rachel Teal, all stroke survivors who took part in the study, and members of the Oxford Translational Neuropsychology Group for contributions to participant recruitment, cognitive test scoring, and data entry, particularly Evangeline Grace Chiu, Romina Basting, and Ellie Slavkova.

