## Supplemental Material for "Domain-specific cognitive impairment 6 months after stroke: the value of early cognitive screening"

**SUPPLEMENTARY MATERIALS**

**SUPPLEMENTARY METHODS:**

Oxford Cognitive Screen considerations:

The OCS is a cognitive screening tool specifically designed to assess common cognitive impairments in patients with stroke and to minimize the impact of stroke on assessment. It takes approximately 15 to 20 minutes to administer. Cognitive assessment was considered feasible if participants were able to complete at least 70% of tasks, allowing for sufficient evaluation of each cognitive domain. Participants who attempted but were unable to complete a task due to an apparent deficit as judged by the assessor were given the minimum score and considered impaired on the task (e.g., in cases of aphasia, picture naming was often scored zero, while those with hemiplegia were able to use their non-dominant arm for the praxis task). In cases where participants were unable to complete a task (e.g., due to time, other administrative constraints, or severe blindness) the untested tasks were not considered when classifying domain impairments (see eTable 3 for information regarding how many patients completed each task).

**SUPPLEMENTARY RESULTS:**

**Supplemental Table 1.** Group demographic differences between those who were followed-up and those who were not.

|  | **Completed 6-month follow-up** | **Did not complete follow-up** | **t-test/Chi^2^-test** |
| --- | --- | --- | --- |
| N | 430 | 436 |  |
| Age at stroke, mean (SD) | 73.86 (12.51) | 74.48 (14.37) | t = 0.677, p = 0.4987 |
| Sex, female N (%) | 200 (46.51) | 205 (56.47) | Chi^2^ = 0.007, p = 0.9352 |
| Education years, mean (SD) | 12.25 (3.55) | 11.62 (2.99) | t = 2.480, p = 0.1323 |
| Handedness, right N (%) | 374 (86.98) | 318 (72.94) | Chi^2^ = 0.000, p = 0.9927 |
| Baseline NIHSS, n (%)* |  |  | Chi^2^ = 5.096, p = 0.0024 |
| NIHSS ≤3 (minor), n (%) | 121 (37.81) | 68 (28.33) |  |
| NIHSS >3 (major), n (%) | 199 (62.19) | 172 (71.67) |  |

*NIHSS was reported in n=320 (74.41%) followed-up and n=240 (55.04%) who weren’t followed-up. Chi2 with Yates correction, two-tailed p-value, p <0.05 considered significant.

**Supplemental Table 2.** Group differences in cognition at acute assessment between those who were followed-up and those who were not. Note correction for multiple comparisons is not reported here as the aim was to just provide general descriptives.

|  | **Completed 6-month follow-up** | **Did not complete follow-up** | **Chi^2^-test** |
| --- | --- | --- | --- |
| N | 430 | 436 |  |
| Language, n (%)  Impaired  Unimpaired | 194/429 (45.2) 235/429 (54.8) | 254/436 (58.3) 182/436 (41.7) | Chi^2^ = 14.20, p = 0.0002 |
| Attention, n (%)  Impaired  Unimpaired | 183/391 (46.8) 208/391 (53.2) | 196/388 (37.1) 192/388 (62.9) | Chi^2^ = 1.07, p =0.3000 |
| Executive function, n (%)  Impaired  Unimpaired | 111/379 (29.3) 268/379 (70.7) | 136/374 (36.4)  238/374 (63.6) | Chi^2^ = 3.96, p =0.0466 |
| Memory, n (%)  Impaired  Unimpaired | 171/429 (39.9) 258/429 (60.1) | 244/436 (56.0) 192/436 (44.0) | Chi^2^ = 21.83, p =0.0001 |
| Number, n (%)  Impaired  Unimpaired | 178/427 (41.7) 249/427 (58.3) | 212/433 (49.0) 221/433 (51.0) | Chi^2^ = 7.19, p = 0.0073 |
| Praxis, n (%)  Impaired  Unimpaired | 112/419 (26.7) 307/419 (73.3) | 167/427 (39.1) 260/427 (60.9) | Chi^2^ = 14.11, p = 0.0002 |
| Unimpaired, n (%) | 7/430 (1.6) | 63/436 (14.5) |  |
| Single domain impairment, n (%) | 107/430 (24.9) | 71/436 (16.3) |  |
| Multi-domain impairment, n (%) | 316/429 (73.7) | 302/436 (69.3) |  |

**Supplemental Table 3.** Cognitive profiles at follow-up in patients impaired and unimpaired acutely.

| **Domain** | **T1 impaired  N (%)** | **T1 impaired and assessed at T2 N** | **Recovered at T2**  **N (%)** | **Persistent impairment at T2 N (%)** | **T1 unimpaired**  **N (%)** | **Unimpaired re-assessed at T2 N** | **Remained unimpaired at T2  N (%)** | **Acquired impairment at T2  N (%)** |
| --- | --- | --- | --- | --- | --- | --- | --- | --- |
| **Language** | **194 (45.2)** | **194** | **89 (45.9)** | **105 (54.1)** | **235 (54.8)** | **235** | **200 (85.1)** | **35 (14.9)** |
| Picture naming | 138 (32.2) | 137 | 78 (56.9) | 59 (43.1) | 291 (67.8) | 291 | 272 (93.5) | 19 (6.5) |
| Semantic understanding | 43 (10.1) | 41 | 39 (95.1) | 2 (4.9) | 385 (90.2) | 369 | 355 (96.2) | 14 (3.8) |
| Sentence reading | 136 (32.5) | 134 | 75 (56.0) | 59 (44.0) | 283 (67.5) | 280 | 254 (90.7) | 26 (9.3) |
| **Attention** | **183 (46.8)** | **178** | **100 (56.2)** | **78 (43.8)** | **208 (53.2)** | **204** | **164 (80.4)** | **40 (19.6)** |
| Egocentric attention | 124 (31.7) | 120 | 90 (75.0) | 30 (25.0) | 267 (68.3) | 262 | 241 (92.0) | 21 (8.0) |
| Allocentric attention | 108 (27.62) | 105 | 71 (67.6) | 34 (32.4) | 283 (72.4) | 277 | 221 (79.8) | 56 (20.2) |
| **Executive Function** | **111 (29.3)** | **106** | **61 (57.5)** | **45 (42.5)** | **268 (70.7)** | **255** | **215 (84.3)** | **40 (15.7)** |
| **Memory** | **171 (39.9)** | **171** | **82 (48.0)** | **89 (52.0)** | **258 (60.1)** | **258** | **210 (81.4)** | **48 (18.6)** |
| Orientation | 90 (21.1) | 89 | 58 (65.2) | 31 (34.8) | 337 (78.9) | 334 | 294 (88.0) | 40 (12.0) |
| Verbal memory | 107 (25.1) | 101 | 63 (62.4) | 38 (37.6) | 320 (74.9) | 313 | 273 (87.2) | 40 (12.8) |
| Episodic memory | 79 (20.2) | 76 | 54 (71.1) | 22 (28.9) | 312 (79.8) | 303 | 277 (91.4) | 26 (8.6) |
| **Number processing** | **178 (41.7)** | **167** | **109 (65.3)** | **58 (34.7)** | **249 (58.3)** | **248** | **221 (89.1)** | **27 (10.9)** |
| Calculations | 73 (17.1) | 64 | 43 (67.2) | 21 (32.8) | 354 (82.9) | 350 | 332 (94.9) | 18 (5.1) |
| Writing | 160 (38.9) | 148 | 106 (71.6) | 42 (28.4) | 251 (61.1) | 247 | 229 (92.7) | 18 (7.3) |
| **Praxis** | **112 (26.7)** | **103** | **73 (70.9)** | **30 (29.1)** | **307 (73.3)** | **291** | **244 (83.8)** | **47 (16.2)** |

Proportion of recovery, persistent impairment, remained unimpaired or acquired a new impairment in those who were assessed both acutely (T1) and at follow-up (T2) in each domain.

**Supplemental Figure 1.** Association between domain subtest impairments at acute assessment, follow-up and from acute to follow-up.

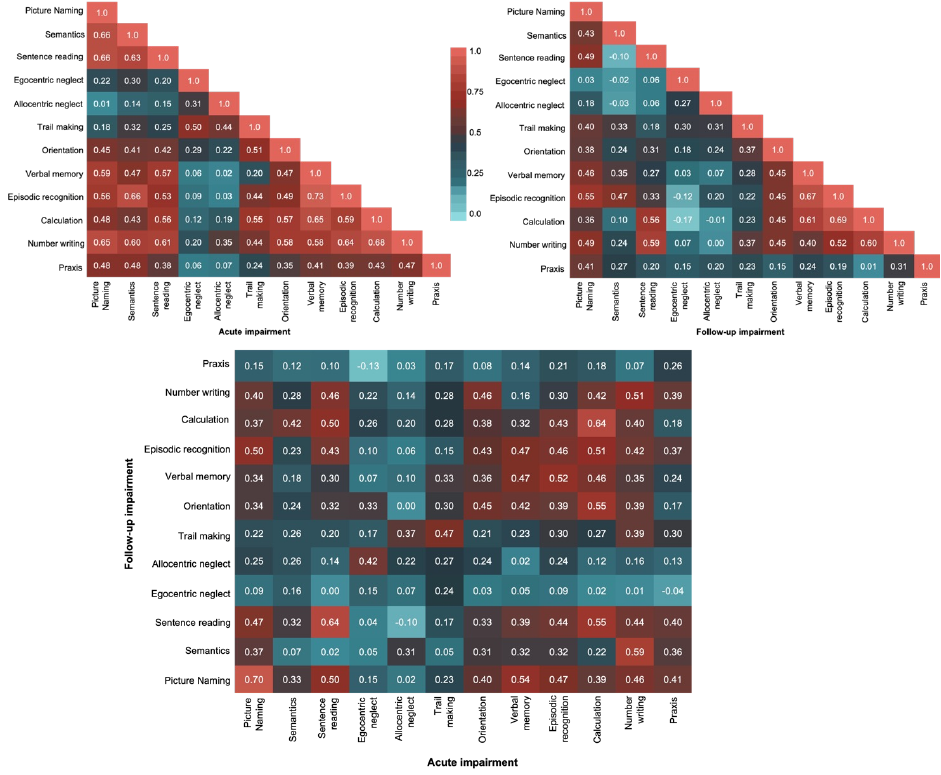

Subtest impairments at acute, follow-up, and acute (x-axis) vs follow-up (y-axis). Tetrachoric correlation coefficient shown in each box with a gradient extending from 0 (light blue) to 1 (light red).

**Supplemental Table 4.** Univariate analyses showing the effect of individual predictor variables on overall cognitive impairment at 6 months.

|  | **β (SE)** | ***t* value** | ***p* value** | **R^2^** | **Adjusted *R*^2^** |
| --- | --- | --- | --- | --- | --- |
| Age | 0.004 (0.001) | 4.090 | 0.000 | 0.046 | 0.044* |
| Education years | -0.013 (0.004) | -3.487 | 0.001 | 0.034 | 0.031* |
| NIHSS | 0.003 (0.003) | 1.118 | 0.264 | 0.004 | 0.001* |
| Days to assessment | 0.001 (0.003) | 0.449 | 0.653 | 0.001 | -0.002 |
| Admission length (days) | 0.001 (0.001) | 1.111 | 0.268 | 0.004 | 0.001* |
|  | **Sum of squares** | **Df** | **Mean square** | **F value** | ***p* value** |
| Sex (male) | 0.026 | 1 | 0.026 | 0.459 | 0.499 |
| Atrial fibrillation | 0.001 | 1 | 0.001 | 0.015 | 0.902 |
| Hypertension | 0.001 | 1 | 0.001 | 0.012 | 0.913 |
| Diabetes | 0.071 | 1 | 0.071 | 1.245 | 0.265 |
| Smoking | 0.129 | 1 | 0.129 | 2.260 | 0.134 |
| Recurrent stroke | 0.332 | 1 | 0.332 | 5.863 | 0.016* |
| Dependence at admission | 0.622 | 1 | 0.622 | 11.14 | 0.001* |
| Charlson Comorbidity Index | 0.187 | 1 | 0.187 | 3.270 | 0.071 |
| * Significance *p* <0.05 | | | |  |  |

**Supplemental Table 5.** Effects of demographic/clinical factors and acute domain-specific impairment on language impairment at 6 months.

| *LANGUAGE* | | | |
| --- | --- | --- | --- |
| **BLOCK 1: Conventional predictors** | β (SE) | *t* value | *p* value |
| Age | 0.004 (0.002) | 2.078 | 0.476 |
| Sex (male) | -0.051 (0.051) | -1.010 | 0.313 |
| Education years | -0.019 (0.007) | -2.629 | 0.009* |
| Atrial fibrillation | -0.022 (0.058) | -0.381 | 0.703 |
| Hypertension | -0.015 (0.051) | -0.288 | 0.774 |
| Diabetes | 0.004 (0.062) | 0.063 | 0.950 |
| Smoking | 0.062 (0.064) | 0.964 | 0.336 |
| NIHSS | 0.009 (0.005) | 1.790 | 0.074 |
| Recurrent stroke | 0.000 (0.053) | -0.008 | 0.994 |
| Days to assessment | 0.003 (0.006) | 0.520 | 0.603 |
| R^2^ = 0.047, Adjusted R^2^ = 0.019, F = 1.649, 10 and 334 df, *p* = 0.09168  AIC 222.9, *p* = 0.3792 | | | |
| **BLOCK 2: Domain-specific impairment** | β (SE) | *t* value | *p* value |
| Age | 0.004 (0.002) | 2.093 | 0.037* |
| Sex (male) | -0.024 (0.047) | -0.516 | 0.606 |
| Education years | -0.008 (0.007) | -1.234 | 0.218 |
| Atrial fibrillation | -0.021 (0.054) | -0.391 | 0.696 |
| Hypertension | -0.003 (0.047) | -0.068 | 0.946 |
| Diabetes | -0.041 (0.059) | -0.701 | 0.483 |
| Smoking | 0.073 (0.059) | 1.230 | 0.220 |
| NIHSS | 0.007 (0.005) | 1.350 | 0.178 |
| Recurrent stroke | -0.004 (0.049) | -0.073 | 0.942 |
| Days to assessment | 0.000 (0.006) | 0.084 | 0.933 |
| **Acute Language** | 0.240 (0.051) | 4.721 | 0.000*^a^ |
| **Acute Attention** | -0.079 (0.048) | -1.638 | 0.102 |
| **Acute Executive** | -0.033 (0.055) | -0.592 | 0.554 |
| **Acute Memory** | 0.194 (0.055) | 3.519 | 0.000*^a^ |
| **Acute Number** | 0.068 (0.057) | 1.203 | 0.230 |
| **Acute Praxis** | 0.101 (0.056) | 1.799 | 0.073 |
| R^2^ = 0.232, Adjusted R^2^ = 0.192, F = 5.863, 16 and 311 df, *p* <0.0001  AIC 196.02, *p* = 0.0004 | | | |
| * Significance *p* <0.05 ^a^ Significant after Bonferroni correction for multiple comparisons | | | |

**Supplemental Table 6.** Effects of demographic/clinical factors and acute domain-specific impairment on attention impairment at 6 months.

| *ATTENTION* | | | |
| --- | --- | --- | --- |
| **BLOCK 1: Conventional predictors** | β (SE) | *t* value | *p* value |
| Age | 0.010 (0.002) | 4.608 | 0.000*^a^ |
| Sex (male) | -0.068 (0.053) | -1.289 | 0.198 |
| Education years | -0.008 (0.008) | -1.010 | 0.313 |
| Atrial fibrillation | -0.100 (0.060) | -1.656 | 0.099 |
| Hypertension | -0.117 (0.053) | -2.207 | 0.028* |
| Diabetes | 0.030 (0.064) | 0.466 | 0.642 |
| Smoking | -0.174 (0.066) | 2.631 | 0.009* |
| NIHSS | 0.009 (0.005) | 1.646 | 0.101 |
| Recurrent stroke | 0.044 (0.055) | 0.799 | 0.425 |
| Days to assessment | -0.001 (0.006) | -0.193 | 0.847 |
| R^2^ = 0.094, Adjusted R^2^ = 0.067, F = 3.447, 10 and 331 df, *p* = 0.0003  AIC 236.3, p=0.0377 | | | |
| **BLOCK 2: Domain-specific impairment** | β (SE) | *t* value | *p* value |
| Age | 0.010 (0.002) | 4.264 | 0.000*^a^ |
| Sex (male) | -0.081 (0.054) | -1.509 | 0.132 |
| Education years | -0.009 (0.008) | -1.126 | 0.261 |
| Atrial fibrillation | -0.113 (0.061) | -1.842 | 0.066 |
| Hypertension | -0.111 (0.054) | -2.043 | 0.042 |
| Diabetes | 0.030 (0.067) | 0.438 | 0.662 |
| Smoking | 0.151 (0.068) | 2.211 | 0.028* |
| NIHSS | 0.005 (0.006) | 0.863 | 0.389 |
| Recurrent stroke | 0.044 (0.056) | 0.788 | 0.431 |
| Days to assessment | -0.002 (0.006) | -0.253 | 0.800 |
| **Acute Language** | -0.035 (0.058) | -0.603 | 0.547 |
| **Acute Attention** | 0.048 (0.055) | 0.864 | 0.388 |
| **Acute Executive** | 0.137 (0.064) | 2.155 | 0.032* |
| **Acute Memory** | 0.059 (0.063) | 0.939 | 0.348 |
| **Acute Number** | 0.020 (0.066) | 0.298 | 0.766 |
| **Acute Praxis** | -0.009 (0.065) | -0.132 | 0.895 |
| R^2^ = 0.128, Adjusted R^2^ = 0.083, F = 2.835, 16 and 309 df, *p* = 0.0003  AIC 235.3, *p*=0.0737 | | | |
| * Significance *p* <0.05 ^a^ Significant after Bonferroni correction for multiple comparisons | | | |

**Supplemental Table 7.** Effects of demographic/clinical factors and acute domain-specific impairment on executive function impairment at 6 months.

| *EXECUTIVE FUNCTION* | | | |
| --- | --- | --- | --- |
| **BLOCK 1: Conventional predictors** | β (SE) | *t* value | *p* value |
| Age | 0.007 (0.002) | 3.544 | 0.000*^a^ |
| Sex (male) | 0.117 (0.046) | 2.557 | 0.011* |
| Education years | -0.005 (0.007) | -0.716 | 0.474 |
| Atrial fibrillation | -0.006 (0.053) | -0.111 | 0.912 |
| Hypertension | -0.059 (0.046) | -1.278 | 0.202 |
| Diabetes | 0.149 (0.056) | 2.678 | 0.008* |
| Smoking | 0.041 (0.058) | 0.714 | 0.476 |
| NIHSS | 0.006 (0.005) | 1.225 | 0.222 |
| Recurrent stroke | 0.061 (0.048) | 1.257 | 0.210 |
| Days to assessment | 0.003 (0.006) | 0.521 | 0.603 |
| R^2^ = 0.09683, Adjusted R^2^ = 0.06978, F = 3.581,10 and 334 df, *p* = 0.00016  AIC 193.44, p=0.0231 | | | |
| **BLOCK 2: Domain-specific impairment** | β (SE) | *t* value | *p* value |
| Age | 0.007 (0.002) | 3.512 | 0.001*^a^ |
| Sex (male) | 0.132 (0.044) | 2.991 | 0.003*^a^ |
| Education years | -0.001 (0.006) | -0.137 | 0.891 |
| Atrial fibrillation | -0.032 (0.050) | -0.644 | 0.520 |
| Hypertension | -0.049 (0.044) | -1.107 | 0.269 |
| Diabetes | 0.098 (0.055) | 1.762 | 0.079 |
| Smoking | 0.046 (0.056) | 0.829 | 0.408 |
| NIHSS | 0.001 (0.005) | 0.154 | 0.878 |
| Recurrent stroke | 0.061 (0.046) | 1.320 | 0.188 |
| Days to assessment | 0.002 (0.005) | 0.425 | 0.671 |
| **Acute Language** | 0.065 (0.048) | 1.363 | 0.174 |
| **Acute Attention** | 0.011 (0.045) | 0.245 | 0.807 |
| **Acute Executive** | 0.225 (0.052) | 4.329 | 0.000*^a^ |
| **Acute Memory** | 0.064 (0.052) | 1.231 | 0.219 |
| **Acute Number** | 0.064 (0.053) | 1.195 | 0.233 |
| **Acute Praxis** | 0.116 (0.053) | 2.020 | 0.028* |
| R^2^ = 0.2274, Adjusted R^2^ = 0.1877, F = 5.721, 16 and 311 df, *p* <0.0001  AIC 170.1, *p*<0.0001 | | | |
| * Significance *p* <0.05 ^a^ Significant after Bonferroni correction for multiple comparisons | | | |

**Supplemental Table 8.** Effects of demographic/clinical factors and acute domain-specific impairment on memory impairment at 6 months.

| *MEMORY* | | | |
| --- | --- | --- | --- |
| **BLOCK 1: Conventional predictors** | β (SE) | *t* value | *p* value |
| Age | 0.004 (0.002) | 1.831 | 0.068 |
| Sex (male) | 0.027 (0.051) | 0.526 | 0.599 |
| Education years | -0.014 (0.007) | -1.899 | 0.058 |
| Atrial fibrillation | -0.038 (0.059) | -0.647 | 0.518 |
| Hypertension | -0.022 (0.052) | -0.430 | 0.667 |
| Diabetes | 0.029 (0.062) | 0.462 | 0.644 |
| Smoking | 0.072 (0.065) | 1.121 | 0.263 |
| NIHSS | 0.002 (0.005) | 0.390 | 0.697 |
| Recurrent stroke | 0.093 (0.054) | 1.728 | 0.085 |
| Days to assessment | 0.010 (0.006) | 1.664 | 0.097 |
| R^2^ = 0.049, Adjusted R^2^ = 0.021, F = 1.721, 10 and 334 df, *p* = 0.07473  AIC 221.4, p=0.379 | | | |
| **BLOCK 2: Domain-specific impairment** | β (SE) | *t* value | *p* value |
| Age | 0.003 (0.205) | 1.237 | 0.217 |
| Sex (male) | 0.061 (0.002) | 1.216 | 0.225 |
| Education years | -0.006 (0.050) | -0.897 | 0.370 |
| Atrial fibrillation | -0.040 (0.057) | -0.707 | 0.480 |
| Hypertension | -0.002 (0.050) | -0.044 | 0.965 |
| Diabetes | -0.033 (0.063) | -0.535 | 0.593 |
| Smoking | 0.071 (0.063) | 1.123 | 0.262 |
| NIHSS | -0.005 (0.005) | -0.864 | 0.388 |
| Recurrent stroke | 0.079 (0.052) | 1.526 | 0.128 |
| Days to assessment | 0.006 (0.006) | 0.969 | 0.333 |
| **Acute Language** | 0.180 (0.054) | 3.530 | 0.001*^a^ |
| **Acute Attention** | 0.043 (0.051) | 0.833 | 0.405 |
| **Acute Executive** | 0.020 (0.059) | 0.338 | 0.735 |
| **Acute Memory** | 0.236 (0.058) | 4.034 | 0.000*^a^ |
| **Acute Number** | 0.020 (0.060) | 0.328 | 0.743 |
| **Acute Praxis** | 0.005 (0.060) | 0.081 | 0.935 |
| R^2^ = 0.171, Adjusted R^2^ = 0.129, F = 4.014, 16 and 311 df, *p* <0.0001  AIC 196.0, *p*=0.0004 | | | |
| * Significance *p* <0.05 ^a^ Significant after Bonferroni correction for multiple comparisons | | | |

**Supplemental Table 9.** Effects of demographic/clinical factors and acute domain-specific impairment on number impairment at 6 months.

| *NUMBER* | | | |
| --- | --- | --- | --- |
| **BLOCK 1: Conventional predictors** | β (SE) | *t* value | *p* value |
| Age | -0.002 (0.002) | -1.277 | 0.203 |
| Sex (male) | 0.052 (0.043) | 1.206 | 0.229 |
| Education years | -0.010 (0.006) | -1.654 | 0.099 |
| Atrial fibrillation | 0.036 (0.050) | 0.717 | 0.474 |
| Hypertension | 0.055 (0.044) | 1.260 | 0.209 |
| Diabetes | -0.030 (0.053) | -0.565 | 0.572 |
| Smoking | 0.044 (0.055) | 0.807 | 0.636 |
| NIHSS | 0.002 (0.004) | 0.473 | 0.420 |
| Recurrent stroke | 0.056 (0.046) | 1.214 | 0.226 |
| Days to assessment | 0.004 (0.005) | 0.811 | 0.418 |
| R^2^ = 0.036, Adjusted R^2^ = 0.007, F = 1.239, 10 and 334 df, *p* = 0.2645  AIC 166.2, p=0.154 | | | |
| **BLOCK 2: Domain-specific impairment** | β (SE) | *t* value | *p* value |
| Age | -0.003 (0.002) | -1.909 | 0.057 |
| Sex (male) | 0.064 (0.042) | 1.520 | 0.130 |
| Education years | -0.004 (0.006) | -0.726 | 0.468 |
| Atrial fibrillation | 0.025 (0.048) | 0.527 | 0.598 |
| Hypertension | 0.055 (0.042) | 1.288 | 0.199 |
| Diabetes | -0.082 (0.053) | -1.553 | 0.121 |
| Smoking | 0.057 (0.053) | 1.070 | 0.286 |
| NIHSS | -0.003 (0.004) | -0.601 | 0.548 |
| Recurrent stroke | 0.043 (0.044) | 0.970 | 0.333 |
| Days to assessment | 0.001 (0.005) | 0.142 | 0.887 |
| **Acute Language** | 0.098 (0.045) | 2.164 | 0.031* |
| **Acute Attention** | 0.052 (0.043) | 1.205 | 0.229 |
| **Acute Executive** | 0.062 (0.050) | 1.259 | 0.209 |
| **Acute Memory** | 0.088 (0.049) | 1.786 | 0.075 |
| **Acute Number** | 0.091 (0.051) | 1.790 | 0.074 |
| **Acute Praxis** | 0.114 (0.050) | 2.260 | 0.025* |
| R^2^ = 0.1549, Adjusted R^2^ = 0.1114, F = 3.561, 16 and 311 df, *p* <0.0001  AIC 157.3, *p*=0.0075 | | | |
| * Significance *p* <0.05 ^a^ Significant after Bonferroni correction for multiple comparisons | | | |

**Supplemental Table 10.** Effects of demographic/clinical factors and acute domain-specific impairment on praxis impairment at 6 months.

| *PRAXIS* | | | |
| --- | --- | --- | --- |
| **BLOCK 1: Conventional predictors** | β (SE) | *t* value | *p* value |
| Age | 0.006 (0.002) | 2.982 | 0.003*^a^ |
| Sex (male) | -0.105 (0.045) | -2.336 | 0.020* |
| Education years | -0.001 (0.007) | -0.201 | 0.841 |
| Atrial fibrillation | 0.002 (0.052) | 0.045 | 0.964 |
| Hypertension | -0.046 (0.045) | -1.025 | 0.306 |
| Diabetes | 0.044 (0.054) | 0.804 | 0.422 |
| Smoking | 0.038 (0.056) | 0.673 | 0.501 |
| NIHSS | 0.004 (0.005) | 0.964 | 0.336 |
| Recurrent stroke | 0.023 (0.0047) | 0.481 | 0.631 |
| Days to assessment | -0.001 (0.005) | -0.199 | 0.842 |
| R^2^ =0.04637, Adjusted R^2^ = 0.01782, F = 1.624, 10 and 334 df, *p* = 0.09827  AIC 209.3, p=0.2818 | | | |
| **BLOCK 2: Domain-specific impairment** | β (SE) | *t* value | *p* value |
| Age | 0.005 (0.002) | 2.606 | 0.010* |
| Sex (male) | -0.097 (0.046) | -2.131 | 0.034* |
| Education years | -0.001 (0.007) | -0.128 | 0.898 |
| Atrial fibrillation | 0.016 (0.052) | 0.315 | 0.753 |
| Hypertension | -0.061 (0.046) | -1.324 | 0.187 |
| Diabetes | 0.006 (0.057) | 0.106 | 0.916 |
| Smoking | 0.059 (0.058) | 1.016 | 0.322 |
| NIHSS | 0.005 (0.005) | 0.992 | 0.311 |
| Recurrent stroke | 0.009 (0.048) | 0.185 | 0.853 |
| Days to assessment | -0.003 (0.005) | -0.504 | 0.614 |
| **Acute Language** | 0.022 (0.049) | 0.447 | 0.655 |
| **Acute Attention** | 0.004 (0.047) | 0.076 | 0.940 |
| **Acute Executive** | -0.004 (0.054) | -0.079 | 0.937 |
| **Acute Memory** | 0.051 (0.053) | 0.954 | 0.341 |
| **Acute Number** | -0.008 (0.055) | -0.147 | 0.883 |
| **Acute Praxis** | 0.189 (0.055) | 3.466 | 0.001*^a^ |
| R^2^ = 0.09153, Adjusted R^2^ = 0.04479, F = 1.958, 16 and 311 df, *p* = 0.01549  AIC 202.9, *p*=0.0746 | | | |
| * Significance *p* <0.05 ^a^ Significant after Bonferroni correction for multiple comparisons | | | |

**Supplementary Analysis (SA):**

As it is plausible that patients with first time stroke events may exhibit a different pattern of cognitive impairment or cognitive recovery, supplementary analyses were run to identify any significant differences in impairment and recovery profile within this group. This sub-sample contained 292 patients who were diagnosed with a first-time stroke event. Table X provides a summary of this sample’s demographic and clinical data alongside statistical comparisons to the full sample. Notably, these characteristics are largely analogous across the full and first-time stroke sample with only proportion independent (corrected alpha = 0.001).

**Supplemental Analysis Table 1:** Cohort demographics and acute clinical characteristics comparisons between all patients and those with first time stroke events. Significant comparisons are starred.

|  | **N = 430** | **N = 288** |
| --- | --- | --- |
| Age at stroke, mean (SD) | 73.86 (12.51) | 72.5 (13.5) |
| Sex (female), n (%) | 200 (46.51) | 142 (48.6) |
| Education years, mean (SD) | 12.25 (3.55) | 12.43 (3.67) |
| Handedness, right n (%) | 374 (86.98) | 252 (86.3) |
| *Stroke subtype, n (%)* |  |  |
| Ischemic | 288 (66.98) | 194 (66.7) |
| Haemorrhagic | 65 (15.12) | 46 (16.0) |
| Mixed | 3 (1.00) | 3 (1.04) |
| Unknown | 74 (17.21) | 49 (0.163) |
| *Lesion side, n (%)* |  |  |
| Left | 153 (35.58) | 101 (35.1) |
| Right | 168 (39.07) | 119 (41.3) |
| Bilateral | 34 (7.91) | 22 (6.94) |
| Undetermined | 75 (17.44) | 50 (16.7) |
| *Major vascular territory, n (%) ^A^* |  |  |
| Anterior | 39 (9.07) | 25 (8.68) |
| Middle | 177 (41.16) | 127 (44.1) |
| Posterior | 59 (13.72) | 41 (13.5) |
| Vertebrobasilar | 55 (12.79) | 36 (12.5) |
| Multifocal | 8 (1.86) | 5 (1.74) |
| Lacunar | 18 (4.19) | 9 (3.13) |
| Undetermined | 74 (17.21) | 49 (16.3) |
| First-ever stroke, n (%) | 292 (67.91) |  |
| NIHSS, median [IQR] ^B^ | 5 [2–10] | 7.35 [ 3-8] |
| NIHSS ≤3, n (%) | 121 (37.81) | 77 (35.6) |
| NIHSS >3, n (%) | 199 (62.19) | 215 (64.3) |
| Modified Rankin Scale, median [IQR] ^C^ | 1 [0–2] | 1 [0-1] |
| Barthel Index, median [IQR] ^C^ | 15 [9–19] | 13 [8-19] |
| Independent at admission, n (%) | 376 (87.44) | **278 (95.1)*** |
| Dependent (care required), n (%) | 54 (12.56) | **14 (4.86)*** |
| *Comorbidities, n (%)* |  |  |
| CCI low (0 – 1) | 268 (62.33) | **292 (100%)*** |
| CCI high (≥2) | 162 (37.67) | **0*** |
| Atrial fibrillation | 109 (25.35) | 65 (21.9) |
| Hypertension | 261 (60.70) | 174 (59.4) |
| Diabetes mellitus | 83 (19.30) | 51 (17.0) |
| *Smoking, n (%)* |  |  |
| Current | 44 (10.23) | 27 (9.38) |
| Past | 34 (7.91) | 26 (9.03) |
| Never | 352 (81.86) | 235 (81.6) |
| Days from stroke to T1 assessment, mean (SD) | 4.38 (4.46) | 4.61 (4.95) |
| Length of stay in hospital, mean days (SD) | 11.64 (11.44) | 11.45 (11.76) |
| CCI: Charlson Comorbidity Index; NIHSS: National Institute of Health Stroke Scale (≤3 minor, >3 major) Patient demographics/clinical characteristics were determined on admission and CCI was scored based on the ICD-9 and ICD-10 scoring scheme through medical records at hospital discharge.  ^A^ Note that vascular supply is a crude characterization of primary territory affected ^B^ NIHSS recorded for n=320 (74%) due to not being routinely recorded in this clinical setting prior to 2014  ^C^ Pre-morbid modified Rankin Scale (mRS) reported for n=248 (58%) ^D^ Barthel Index was reported for n=141 (33%) ^E^ Dependence was categorized as requiring formal support (by family n=18, by carer n=36) | | |

***Prevalence of and associations between domain-specific impairments acutely and at 6 months***

Within the subsample of patients with first-time stroke diagnoses, overall prevalence of post-stroke cognitive impairment acutely and at 6 months is shown in SA Table 2 below. Deficit prevalence which are significantly different from the full sample are denoted with stars (chi-squared tests, Bonferroni-corrected alpha level = 0.00125). All other comparisons are not significant, indicating a similar cognitive profile in the first stroke and full cohort groups.

**Supplemental Analysis Table 2.** Prevalence of post-stroke cognitive impairments acutely and at 6 months.

| **Domain** | **Assessed acute  (N)** | **Acute impaired  N (%)** | **Assessed follow-up N** | **Follow-up impaired  N (%)** |
| --- | --- | --- | --- | --- |
| **Language** | **292** | **134 (45.7)** | **292** | **93 (31.8)** |
| Picture naming | 292 | 98 (33.4) | 291 | 49 (16.8) |
| Semantic understanding | 291 | 30 (10.3) | 282 | 11 (3.90) |
| Sentence reading | 287 | 96 (33.3) | 286 | 63 (22.0) |
| **Spatial attention** | **268** | **124 (46.1)** | **281** | **103 (36.7)** |
| Egocentric attention | 268 | 89 (33.1) | 281 | 65 (23.1) |
| Allocentric attention | 268 | 69 (25.7) | 281 | 60 (21.4) |
| **Executive Function** | **257** | **76 (29.5)** | **274** | **61 (22.3)** |
| **Memory** | **291** | **110 (37.7)** | **292** | **87 (29.8)** |
| Orientation | 289 | 56 (19.3) | 290 | 48 (16.6) |
| Verbal memory | 290 | 72 (24.7) | 284 | 48 (16.9) |
| Episodic memory | 265 | 50 (18.8) | 283 | 33 (11.7) |
| **Number processing** | **289** | **122 (42.1)** | **285** | **56 (19.6)** |
| Calculations | 289 | 50 (17.2) | 284 | 22 (7.75) |
| Writing | 280 | 110 (39.1) | 280 | 49 (17.5) |
| **Praxis** | 285 | 79 (27.6) | 271 | 48 (17.7) |
| **Any domain** | 292 | 238 (81.5)** | 292 | 200 (68.5) |
| **Multi-domain** | 292 | 178 (61.0)** | 292 | 134 (45.9) |
| **Single domain** | 292 | 64 (21.9) | 292 | 66 (22.6) |

***Predictors of proportion of impaired cognitive domain subtests at 6 months***
Hierarchical regression was employed to identify acute clinical and cognitive factors predicting cognitive performance at follow-up assessment within patients with first-time stroke events. In the base model (Block 1), acute common risk factors significantly predicted proportion of subtests impaired at follow up within first-time stroke survivors (*F*(10,218)=3.06, *p*<0.0001, adjusted *R*^2^=0.083).In this model, age (*p*<0.0002), education (*p*=0.02), smoking (*p*=0.029) and lesion volume (*p*=0.016) were significant predictors, though only age remained significant after correction (Bonferroni corrected alpha level=0.005). Proportion of acute subtests impaired was then added to this base model (Model 1, Block 2), which, as in the full sample, markedly improved the model, adjusted *R*^2^=0.257 (*p*<0.0001). These results agree well with findings from the full considered cohort. After correction for multiple comparisons, only age (*p*<0.0001) and acute cognition (proportion of impaired subtests acutely) (*p*<0.0001) remained significant. These results are identical to those found in the full sample. Finally, acute domain-specific cognitive impairments were added to the conventional predictor base model (Model 2, Block 2), which again improved the model fit, adjusted *R*^2^=0.298 (*p*<0.0001). As found in the full sample analysis, acute language (*p*=0.0009), memory (p = 0.0004) and age (*p*=0.001) remained significant after correction.

***Predictors of domain-specific cognition at 6 months***

Subsequently, a series of 6 hierarchical regression analyses were conducted to identify acute clinical and cognitive factors predictive of individual domain impairments at follow-up within the first-time stroke subsample. For each of these models, a base model reported how well conventional predictors explained the occurrence of domain-specific chronic impairment while an expanded model investigated how the addition of all acute cognitive domain data improved this fit. As found within the full sample, the fit of all considered models improved following the addition of cognitive data.

In the language domain, the addition of all acute cognitive domains to conventional predictors improved the model from AIC 222.9 (p= 0.379) to AIC 221.4 (p= 0.0048) (eTable 5). Language impairment was best predicted by acute language (OR = 2.8265, *p*=0.0149), memory (OR= 2.6071, *p=* 0.0388), and age (OR= 1.0433, p = 0.0301), but these effects did not survive correction for multiple comparisons. Similarly, the base model predicting attention improved from adjusted AIC 236.27 (p= 0.0377) to adjusted AIC 235.25 (p= 0.0757) with the addition of acute cognitive function. The only predictor surviving multiple corrections was found to be stroke age (OR= 1.0673, *p=0.0006*).The model predicting executive function improved from AIC 193.44 (p= 0.023) to AIC 170.07 (p < 0.001) The strongest predictor of executive function impairment at follow-up was an acute executive impairment (OR= 6.2655, *p=* 0.00056) and age (OR = 1.0782, p = 0.0029 ).The model predicting memory at 6 months improved from AIC 221.36 (p= 0.270) to AIC 218.17 (p< 0.001) with the addition of acute domain cognitive function to the base model. The strongest predictors of memory impairment at follow-up were acute language (OR= 1.1919, *p=* 0.0189). The regression predicting number impairment improved from AIC 166.2 (p= 0.1536) to adjusted AIC 157.3 (p= 0.007). As in the full sample, no individual predictors remained significant after correction. Lastly, praxis impairment at follow-up improved from AIC 209.34 (p= 0.2818) to adjusted AIC 202.88 (p= 0.07464), with acute praxis (OR= 3.2656, *p=* 0.00798) significantly predicting chronic praxis impairment.

Overall, these results agree well with those found within the full sample. This suggests that the predictive relationships identified in this study are common across all stroke survivors, regardless of whether they had a single event or multiple stroke events.
